# Functional and Structural Characterization of LRRK2 p.V1447L in Parkinson’s Disease

**DOI:** 10.1101/2025.04.22.25326118

**Authors:** Neringa Pratuseviciute, Pawel Lis, Sacha Weber, Nicolas Gruchy, Lionel Arnaud, Dario R Alessi, Esther Sammler

**Affiliations:** Medical Research Council Protein Phosphorylation and Ubiquitylation Unit, University of Dundee, Dundee, United Kingdom; Institut du Cerveau-Paris Brain Institute ICM, Sorbonne Université, Inserm 1127, CNRS 7225, Hôpital de la Pitié Salpêtrière Paris, Paris, France; Université Caen Normandie, Normandie Univ, Biotargen UR7450, CHU de Caen, Service de génétique, 14000 Caen France; Département de Génétique Médicale, APHP Sorbonne Université, Paris, France; Division of Neuroscience, School of Medicine, University of Dundee, Dundee, United Kingdom

**Author notes:** **Correspondence to:** Dr. Esther Sammler, MRC Protein Phosphorylation and Ubiquitylation Unit, School of Life Sciences, Dundee DD1 5EH, UK.

**Keywords:** LRRK2, Parkinson’s disease, genetics, peripheral blood neutrophils, Rab10 phosphorylation

## Abstract

**Background:** Gain-of-kinase-function variants in LRRK2 are a leading cause for Parkinson’s disease (PD).

**Objectives:** We tested the functional impact of a novel LRRK2 variant p.V1447L identified in a young onset PD patient *in vivo* in peripheral blood as well as in a robust cellular assay in addition to other variants in close proximity to V1447.

**Methods:** We measured LRRK2-dependent Rab10 phosphorylation in neutrophils and monocytes of a LRRK2 p.V1447L carrier with PD. We deployed structural modelling and evaluated the impact of additional LRRK2 variants at and around LRRK2 V1447.

**Results:** LRRK2 p.V1447L strongly activates LRRK2 kinase activity. We identified additional variants in the LRRK2 ROC:COR_B_ interface with critical impact on kinase activity and show that different substitutions at the same residue can have opposing effects.

**Conclusions:** We recommend reclassifying LRRK2 p.V1447L from VUS to likely pathogenic. Our study expands the range of putative Loss-of-kinase function variants to LRRK2 missense variants.

---

The global increase in people living with Parkinson’s disease (PD), now over 10 million cases, highlights the urgent need for disease modifying treatments^1^. Thus, the outcome of clinical trials, especially those targeting molecular mechanisms of genes that have been linked to causing or significantly increasing the risk for PD, are eagerly awaited. The Leucine rich repeat kinase 2 (LRRK2) is a high value target for disease modification in PD with clinical trials currently underway^2, 3^. Heterozygous gain-of-kinase function variants in *LRRK2* represent the most frequent cause of monogenetic PD accounting for 1-4% of all PD cases worldwide, but this data is mainly based on a few of the more common variants such as p.G2019S and p.R1441G^4–6^. Given the sheer number of rare *LRRK2* variants of unknown clinical significance (VUS) reported thus far, it is important to determine their functional impact and pathogenicity^7, 8^.This will have implications for PD patient stratification, support genetic counselling and possibly predict response to treatment with LRRK2 therapeutics.

Here we report on the functional analysis of a novel LRRK2 VUS (p.V1447L) identified in a sporadic PD patient with early disease onset. The V1447 residue is located within a region of the GTPase domain of LRRK2 known to play an important role in controlling kinase activity. We demonstrate that this variant significantly activates LRRK2 kinase activity *in vivo* in patient peripheral blood as well as in a robust cellular LRRK2 overexpression assay. Our structural analysis indicates that V1447L participates in a network of interactions involving at least 6 other residues including R1441 and Y1699 that are linked to *LRRK2* pathogenic mutations and that the V1447L variant stimulates LRRK2 kinase activity by destabilising the inactive conformation of LRRK2 in the same manner as mutations on other residues in this region. Based on our data, we recommend reclassification of LRRK2 p.V1447L from VUS to at least likely pathogenic.

## Materials and methods

For clinical sample analysis 40ml of fresh blood was collected from the patient and an unrelated control for neutrophil and monocyte isolations and *ex vivo* treatment with and without the specific LRRK2 kinase inhibitor MLi-2 (200nM, 30min)^9^ prior to cell lysis as before ^10, 11^. For extended methods on isolations, HEK293 cell transfections and quantitative immunoblot analysis refer to Supplementary Data.

## Results

### Index patient and genetic testing

The patient is a Caucasian woman in her early fifties, with a diagnosis of PD established before the age of 45. The patient has no known family history of PD or tremors. Because of the relatively early age at PD onset, along with some atypical clinical features, genetic testing via next-generation sequencing (NGS) was performed. This did not reveal a pathogenic variant or relevant copy number variation in any of the common PD associated genes, instead, a rare heterozygous LRRK2 VUS (c.4339G>C, p.V1447L) and carrier status for a common GBA1 PD risk variant, p.T408M (c.1223 C>T), were identified.

To our knowledge, the LRRK2 p.V1447L variant has not been reported in PD patients before, at least not in the literature, the MDSGene database (https://www.mdsgene.org) or the PDvariant browser (https://pdgenetics.shinyapps.io/VariantBrowser/)^12, 13^. In the gnomAD database (v4.1.0), LRRK2 p.V1447L is also absent ^14^. However, MDSGene lists 6 PD patients where valine at this residue is substituted with methionine, and this was previously found to activate LRRK2 kinase activity in a robust cellular assay ^15^. LRRK2 p.V1447M is generally rare and not found in gnomAD. At the same residue, other substitutions have been reported in gnomAD including LRRK2 p.V1447E and p.V1447V variants (synonymous) that were both found once and the p.V1447G variant that was found 18 times in the heterozygous state. These variants have not been reported in PD patients in the databases that we interrogated, and no functional data exists.

### Elevated LRRK2 kinase activity in peripheral blood of the LRRK2 p.V1447L carrier with PD

We next explored whether the PD patient carrying the LRRK2 p.V1447L variant had increased LRRK2 kinase pathway activity *in vivo* in peripheral blood. Using LRRK2-dependent Rab10 substrate phosphorylation as a readout for LRRK2 kinase activity^10, 16^, we observed an over 3-fold activation in both peripheral blood neutrophils and monocytes in comparison to a healthy control (Fig.1 A&B). Near complete dephosphorylation of pThr73-Rab10 was observed in response to LRRK2 kinase inhibitor treatment with MLi-2, confirming that measured Rab10 phosphorylation was mediated by the LRRK2 kinase.

**Figure 1.**
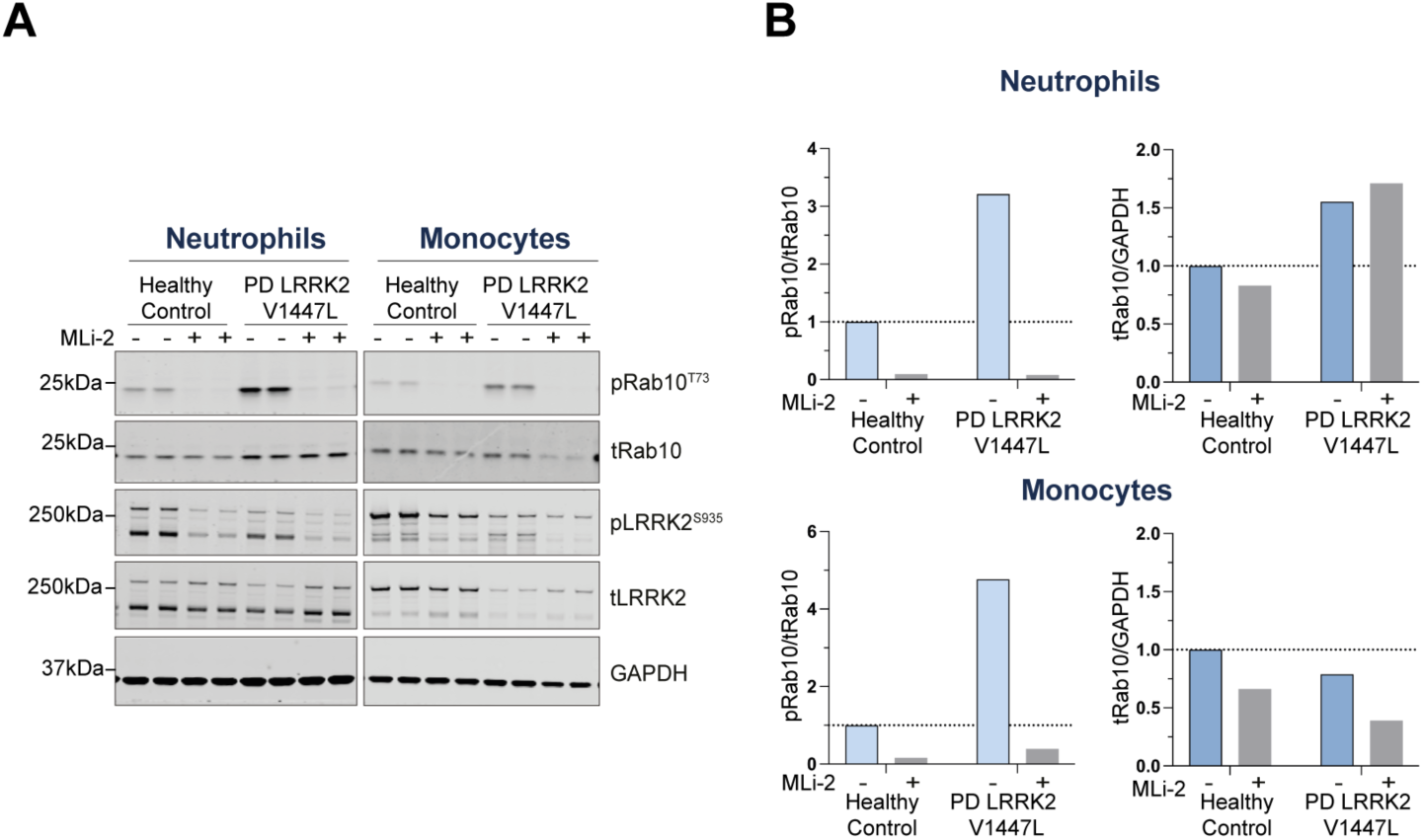
LRRK2 V1447L variant increases LRRK2 dependent Rab10^Thr73^ phosphorylation in PD patient-derived clinical samples. (A) Neutrophils and monocytes were isolated from a PD patient carrying LRRK2 p.V1447L variant and an age-matched healthy control. Samples were treated *ex vivo* with or without 200nM MLi-2 for 30min and 10ug of each sample was loaded. Following Western Blot analysis, membranes were incubated with indicated antibodies and imaged using LICOR Odyssey CLx imaging system. Quantified results were normalized to the healthy control and expressed as (B) pRab10/total Rab10 and total Rab10/GAPDH.

### Structural modelling of the LRRK2 p.V1447L variant

Analysis of the high resolution Cryo-EM structure of full length inactive LRRK2 (PDB, 7LI4)^17^ reveals that the V1447 residue is located within the ROC domain, lying within a beta-sheet region that interacts with an alpha-helix on the surface of the ROC domain, encompassing residues 1424-1442, termed the α3-helix^18^ (Fig. 2A). The V1447 residue is highly conserved amongst species with a Consurf score of 9/9^19^, suggesting that it plays a functionally critical role. The α3-helix that the V1447 residue interacts with encompasses the previously characterised p.A1440P and p.R1441/C/G/S LRRK2 kinase activating pathogenic variants (Fig 2A). The surface of the α3-helix opposite to V1447 interacts with the COR_B_ domain (Fig 2A). Residues in this COR_B_ domain interface that interact with the ROC domain α3-helix include previously characterized LRRK2 kinase activating variants p.Y1699C^4^ and p.F1700L^20^. The data suggest that the network of interactions between the ROC beta-sheet, α3-helix and COR_B_ plays a critical role in stabilising the inactive from of LRRK2. Previous work has revealed that disrupting the ROC-COR_B_ interface by introduction of the aforementioned pathogenic variants promotes LRRK2 activation^15, 17, 21^. Our data suggest that the p.V1447L and p.V1447M variants in the beta-sheet region impact the positioning and/or stability of the α3-helix. This is predicted to increase LRRK2 kinase activity by destabilizing its inactive conformation by impacting the ROC-COR_B_ interface (Fig 2A).

**Figure 2.**
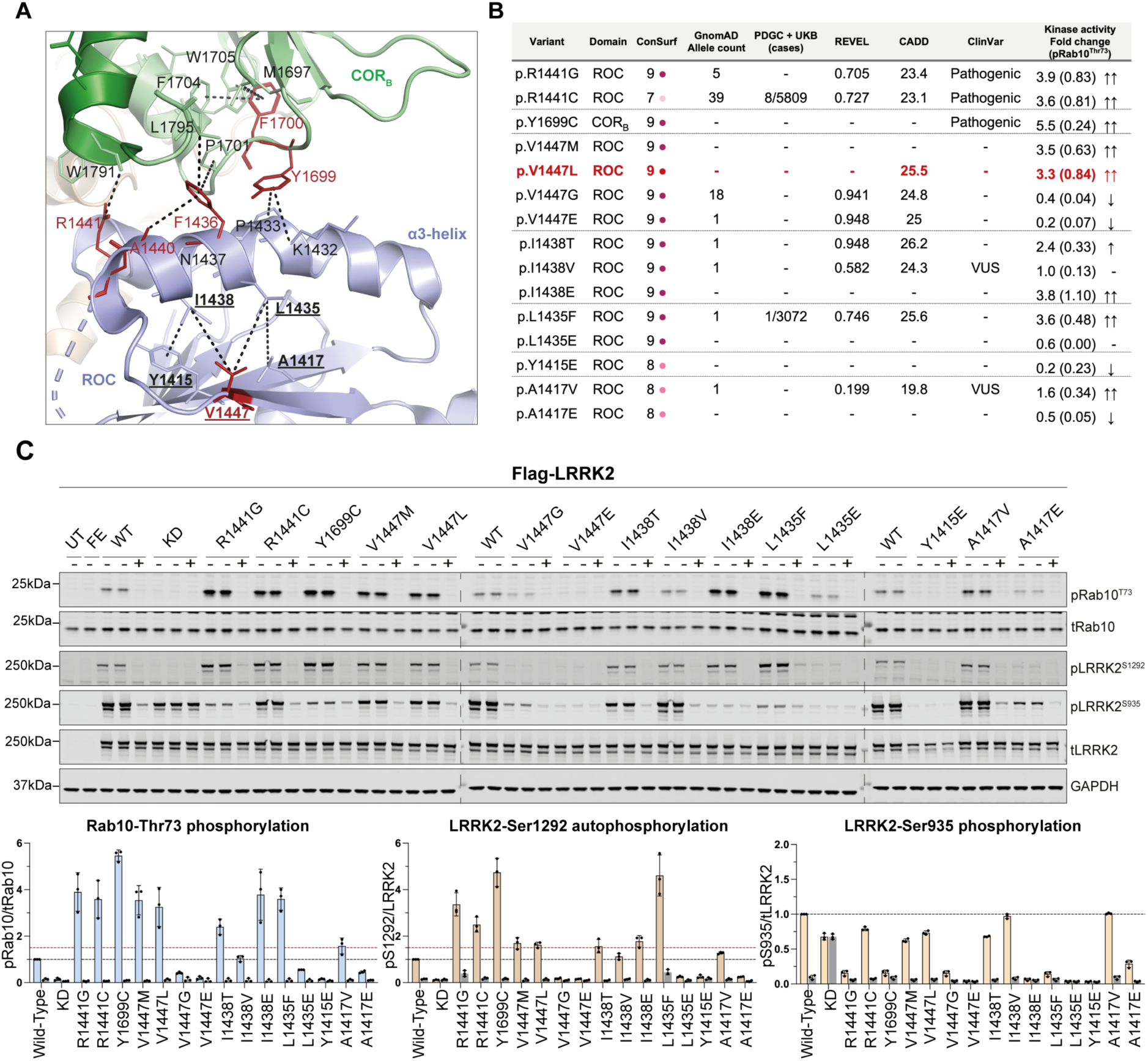
Structural and functional analysis of LRRK2 variants at the ROC:COR_B_ interdomain interface identifies several novel kinase activating variants. (A) Structural representation of the ROC:COR_B_ interdomain interface, obtained using Cryo-EM structure of full length inactive LRRK2 (PDB, 7LI4)^17^. Indicated in red are residues where pathogenic LRRK2 activating variants have been identified. (B) Table representing the evolutionary conservation and frequency of LRRK2 variants in general healthy population and PD patients. REVEL and CADD scores provide *in silico* predictions regarding the probability that the variants may be pathogenic. Finally, the table summarizes the findings from the cellular assay of the LRRK2 variant analysis. Fold change >3-fold indicates that variant **i**s **strongly activating** (↑↑) while <3 but >1.5-fold, **moderately activating** (↑). Conversely, a downward arrow (↓) indicates that pRab10 levels are <0.5 relative to LRRK2 wild-type, therefore **inactivating**. The quantified results are representative of average of three biological replicates. (C) Functional analysis of new variants within ROC:COR_B_ interdomain interface using overexpression assay in HEK293 cells with wild-type LRRK2 (WT) and kinase dead (KD=D2017A) as positive and negative controls respectively. 20ug of cell lysates were subjected to multiplexed immunoblot analysis, quantified results were then normalized to the LRRK2 wild-type and expressed as pRab10/total Rab10, pSer935/total LRRK2 and pSer1292/total LRRK2. Red dotted line at y=1.5 emphasizes phosphorylation levels above 1.5-fold relative to the LRRK2 wild-type. Each data point represents one biological replicate.

### Functional analysis of variants at and around LRRK2 V1447 and ROC:COR_B_ interface

To further explore this hypothesis, we attempted to destabilize this interface by studying the impact of mutations into other residues located at the beta-sheet:α3-helix interface. We focused on four residues namely Y1415 and Y1417 (located on the beta-sheet interface) as well as L1435 and I1438 (located on the α3-helix interface). Reviewing accessible PD databases as well as gnomAD we found 6 variants associated with these residues namely p.V1447G, p.V1447E, p.I1438T, p.I1438V, p.L1435F and p.A1417V (Fig. 2B), and tested the impact of these variants on LRRK2 kinase activity using our HEK293 cell assay^15^. Additionally, we tested the functional impact of the p.I1438E, p.L1435E, p.Y1415E, p.A1417E variants, not reported in databases, on LRRK2 activity. These residues are highly conserved with a Consurf score of 8-9 (Fig. 2B).

The novel p.V1447L variant identified in our sporadic PD patient, along with the previously reported p.V1447M, strongly activated LRRK2 kinase activity with a 3.5-fold increase in LRRK2-dependent Rab10 substrate phosphorylation and a moderate enhancement of Ser1292 LRRK2 autophosphorylation (1.5-fold) compared to LRRK2 wild-type. LRRK2 biomarker site Ser935 phosphorylation was marginally reduced. In contrast, mutating the V1447 residue to glycine (p.V1447G) or glutamic acid (p.V1447E) reduced Rab10 phosphorylation and abolished Ser1292 as well as Ser935 LRRK2 phosphorylation (Fig. 2C). For the I1438 residue, substitutions with glutamic acid (p.I1438E) and threonine (p.I1438T) increased Rab10 phosphorylation 2- and 4-fold respectively and enhanced Ser1292 autophosphorylation (1.5-fold), while only p.I1438T reduced Ser935 phosphorylation. In contrast, the I1438V variant had no impact on either LRRK2 kinase activity or Ser935 phosphorylation. The p.I1438E variant has not been reported in PD patients to our knowledge, while the p.I1438T variant has been reported once in the heterozygous state in a Finnish individual in gnomAD but not in the PD variant browser^14^. A nearby L1435F variant (located within the α3-helix), was reported in a PD patient^13^ and once in a South Asian individual in gnomAD and stimulated LRRK2 kinase pathway ∼ 4-fold as measured by Rab10 phosphorylation. On the other hand, when the same residue was mutated to glutamic acid (p.I1435E), a missense variant not reported in humans, LRRK2 kinase activity was reduced. Similarly, the p.A1417E variant, not reported in humans, decreased LRRK2 activity, whereas p.A1417V, observed once in a European individual in gnomAD, showed mild activation in LRRK2 substrate phosphorylation (1.5-fold) but no effect on Ser1292 auto- and Ser935 phosphorylation.

## Discussion

We describe a sporadic PD patient with disease onset in her early forties carrying a rare heterozygous LRRK2 p.V1447L variant of unknown clinical significance as well as a GBA1 risk variant for PD (p.T408M). While her GBA1 variant carrier status could potentially account for the relatively early disease onset, we were interested to explore the functional impact of the LRRK2 p.V1447L variant. We found substantial LRRK2 kinase pathway activation with Rab10 substrate phosphorylation as a readout *in vivo* in peripheral blood neutrophils and monocytes derived from the patient compared to an unrelated control (Fig 1). This was confirmed in our robust LRRK2 overexpression system in HEK293 cells where the p.V1447L variant resulted in an over 3-fold increase in LRRK2 kinase activity. This effect size is in the range of pathogenic LRRK2 R1441 hotspot variants including p.R1441G/C (Fig. 2C) and much higher than for the common LRRK2 p.G2019S variant (around 1.5-fold)^15, 22^. Given that the LRRK2 p.V1447L variant is very rare and that we show clear *in vivo* functional data that this variant hyperactivates LRRK2 kinase activity, we recommend the reclassification of LRRK2 p.V1447L from VUS to likely pathogenic for PD in line with ACMG guidance^23^.

Previous work^17^ highlighted the importance of the structural interplay between the ROC:COR_B_ interface given that it is a hotspot for some of the most activating pathogenic LRRK2 variants (including p. A1440P^24^, p.R1441G^25^/C^4^/H^25, 26^ or p.Y1699C^4^ and p.F1700L^20^). While V1447 does not directly map into the ROC:COR_B_ interface, we hypothesized that it is part of a small hydrophobic area that interacts and likely helps position or stabilize the α3-helix so it can optimally interact with the COR_B_ interface to stabilize the inactive conformation of LRRK2. Indeed, a network of interactions can be mapped from the ROC beta-sheet domain that the V1447 residue is located on to the α3-helix and the COR_B_ domain, that encompasses many LRRK2 activating variants. We have also identified another α3-helix residue that interacts with the ROC beta-sheet proximal to V1447 namely I1438 that when mutated to glutamic acid or threonine also leads to substantial activation of LRRK2. The available data indicate that variants that disrupt this network of interactions promote LRRK2 activation by destabilising the inactive conformation of LRRK2. It should be noted that several mutations we generated in this region namely p.Y1415E and p.A1417E in addition to p.V1447G and p.V1447E inhibited LRRK2 activity. It is possible that these mutations further stabilize the inactive conformation and/or have other structural effects that block the catalytic LRRK2 kinase activity. We propose that the functional impact of amino acid substitutions in this region strongly affects LRRK2 kinase activity in a manner that is influenced by the degree of stabilization of the inactive conformation of LRRK2.

Our work emphasises the importance of performing functional studies in parallel with analysis of the LRRK2 structure to assess how different variants, including different substitutions at the same residue, affect LRRK2 kinase activity, as a key component of the framework for evaluating the pathogenicity of missense variants in LRRK2. If a LRRK2 substitution results in LRRK2 kinase activation in a cellular assay and this can be confirmed *in vivo* in human blood, this provides strong consensus that LRRK2 variants are pathogenic and exert their disease promoting effects by activating LRRK2 kinase activity. Variants that reduce LRRK2 activity are not expected to have any adverse health effects at least in the heterozygous state, as humans with heterozygous loss-of-function LRRK2 variants are reported to be healthy^27^. Interestingly, our study also expands the range of putative loss-of-kinase function variants in LRRK2, to LRRK2 missense variants. In summary, as more and more PD patients will have their genomes sequenced and with the emerging potential of LRRK2 kinase inhibitor disease modifying treatments^3^, it will become increasingly more important to annotate the pathogenicity of a large number of LRRK2 variants of unknown clinical significance. Patients carrying LRRK2 activating variants are expected to be the most likely candidates to benefit from LRRK2 inhibitor therapy. Based on current evidence, we also recommend reclassifying the LRRK2 p.V184L variant from a variant of uncertain significance (VUS) to at least “likely pathogenic,” and suggest that individuals harbouring this variant be considered eligible for LRRK2 inhibitor treatment.

## Acknowledgments

We sincerely thank the patient and volunteer for their generous donation of the blood samples as their contribution to the study has been invaluable. We would also like to thank the technical support of the MRC Protein Phosphorylation and Ubiquitylation Unit Reagents and Services at the University of Dundee with special acknowledgment to Melanie Wightman for her work in generating the plasmids utilized in this study.

## Data availability

Data that support the findings of this study are available from the corresponding author, upon reasonable request.

## Author Contributions

1. Research project: **A.** Conception **B.** Organization **C.** Execution
2. Statistical Analysis: **A.** Design **B.** Execution **C.** Review and Critique
3. Manuscript Preparation: **A.** Writing of the first draft **B.** Review and Critique

NP: 1A, 1B, 1C, 2A, 2B, 3ª

PL: 1A, 1C, 3A, 3B

SW: 2C, 3B

NG: 2C, 3B

LA: 1A, 2C, 3B

DRA: 1A, 3B

ES: 1A, 1B, 2A, 3A

## Ethics Statement

Study participants have provided written informed consent, and ethical approval has been granted by the committee of the University Hospital Centre of Caen, France.

## Peripheral blood collection and neutrophil & monocyte isolation

For the analysis of LRRK2 kinase pathway activity in human samples, 40ml of fresh blood was collected via venesection from the patient and an unrelated aged-matched healthy control for immediate peripheral blood neutrophil and monocyte isolations via immunomagnetic negative selection, consistent with previous methods^1^. Prior to lysis, the cells were treated *ex vivo* with and without the specific LRRK2 kinase inhibitor MLi-2 (200nM, 30min), synthesized by Natalia Shpiro (University of Dundee).

## Plasmids, cell culture and transient transfection in HEK293 cells

All plasmids were obtained from the MRC PPU Reagents and Services (https://mrcppureagents.dundee.ac.uk) (see Table 1). HEK293 overexpression assay was performed as described previously^2^. Briefly, HEK293 cells were grown in DMEM (Dulbecco s Modified Eagle Medium) supplemented with 10% (v/v) foetal calf serum (FBS), 2 mM L-glutamine, 100 U/ml penicillin. The cells were seeded in 6-well plates and at 80-90% confluency transfected with 2ug of each of the Flag-LRRK2 plasmids, including Flag-Empty control plasmid using polyethylenimine (PEI) transfection reagent (1:3 DNA:PEI ratio) all in 300uL Opti-MEM (Gibco). 16-20 hours after transfections the cells were treated with either DMSO or LRRK2 kinase inhibitor MLi-2 (200nM, 90min) and lysed with ice cold 1% (v/v) Triton lysis buffer. The cell debris was cleared by centrifuging the samples at 17 000g for 15min at 4°C and protein concentration determined by BCA protein assay (Thermo Fisher Scientific Cat #23225).

**Table 1:** Plasmids generated and used in this study:

| **DU NUMBER** | **CONSTRUCT** | **PLASMID** |
| --- | --- | --- |
| **DU41799** | Flag Empty | pCMV5 |
| **DU6841** | Flag LRRK2 Wild-type | pCMV5 |
| **DU80014** | Flag LRRK2 Y1415E | pCMV5 |
| **DU77993** | Flag LRRK2 A1417V | pCMV5 |
| **DU77994** | Flag LRRK2 A1417E | pCMV5 |
| **DU77991** | Flag LRRK2 L1435F | pCMV5 |
| **DU77992** | Flag LRRK2 L1435E | pCMV5 |
| **DU77990** | Flag LRRK2 I1438T | pCMV5 |
| **DU80013** | Flag LRRK2 I1438V | pCMV5D |
| **DU77995** | Flag LRRK2 I1438E | pCMV5 |
| **DU26477** | Flag LRRK2 R1441G | pCMV5 |
| **DU13078** | Flag LRRK2 R1441C | pCMV5 |
| **DU62501** | Flag LRRK2 V1447M | pCMV5 |
| **DU77444** | Flag LRRK2 V1447L | pCMV5 |
| **DU77977** | Flag LRRK2 V1447G | pCMV5 |
| **DU77981** | Flag LRRK2 V1447E | pCMV5 |
| **DU26486** | Flag LRRK2 Y1699C | pCMV5 |
| **DU10128** | Flag LRRK2 D2017A (Kinase dead) | pCMV5 |

## Quantitative immunoblot analysis

Cell lysates were prepared at a concentration of 2µg/µL in NuPage LDS Sample Buffer (x4) with 5% β-mercaptoethanol and boiled at 96°C for 10min. 10ug or 20ug of each sample was loaded on the NuPAGE Bis-Tris 4-12% gradient gels and electrophorized at 100V for ∼2h. Using a nitrocellulose membrane, the gels were transferred at 90V for 90min in 1X Transfer Buffer (48mM Tris-HCl and 39mM glycine) and blocked for 30min in 5% skim dry milk diluted in TBS-T. The membranes were left to incubate overnight at 4°C with primary antibodies diluted at 1 µg/ml: multiplexed monoclonal anti-LRRK2 mouse (NeuroMab #75-253) and anti-pS935 LRRK2 rabbit (Abcam #ab133450) and multiplexed monoclonal anti-Rab10 mouse (Nanotools #0680-100) and anti-MJFF-pRab10 rabbit (Abcam #ab230261) antibodies. LRRK2 autophosphorylation site was visualized using monoclonal anti-pS1292 LRRK2 rabbit (Abcam #ab203181) at 1:1000 dilution. GAPDH (Santa Cruz #sc-32233) was used as a loading control at 1:5000. After 30min wash, the membranes were incubated 1h at room-temperature with multiplexed fluorescent secondary antibodies: 1:10000 goat anti-mouse IRDye 680LT and 1:10000 goat anti-rabbit IRDye 800CW (LI-COR). Finally, the membranes were rinsed, and the signals acquired with LI-COR Odyssey CLx imaging system. Post data analysis the results were visualized using GraphPad Prism V10.0.3.

## Raw Data

### Raw Western Blot files

Comprehensive Western Blot files visualizing the raw images as acquired from the LICOR Odyssey CLx. The images are provided for each figure, along with additional biological replicates associated with Figure 2C that are not included in the primary manuscript.

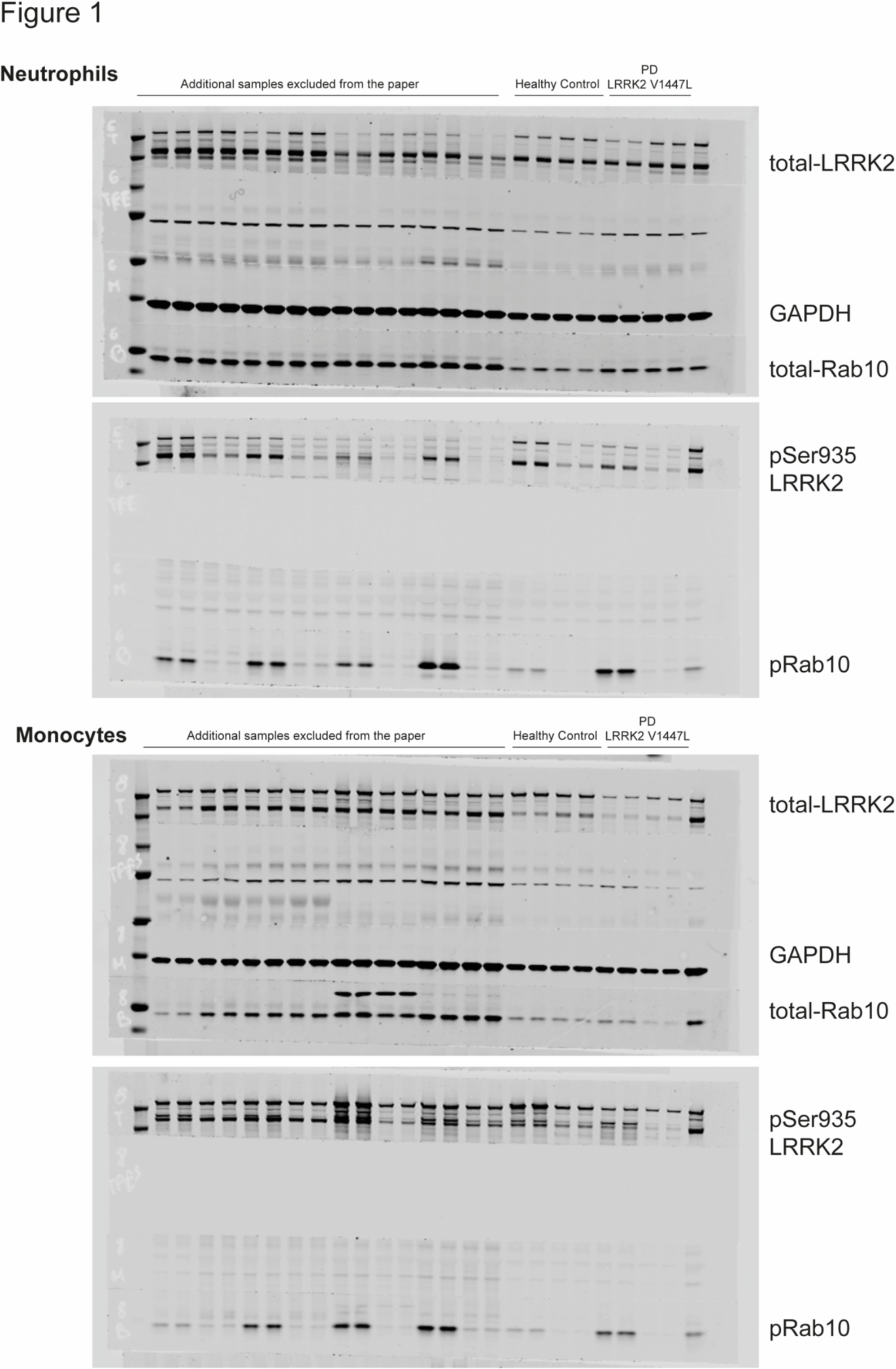

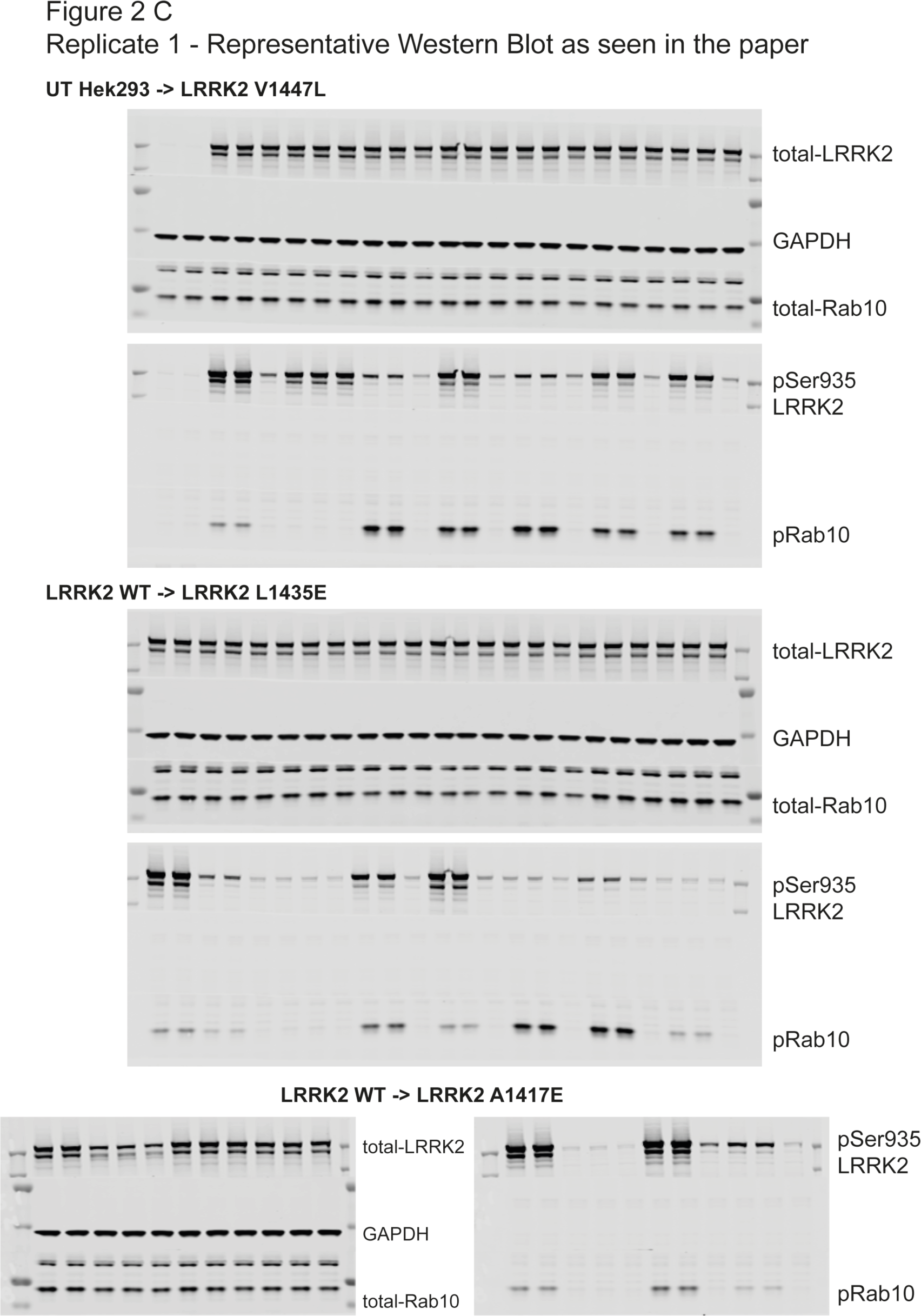
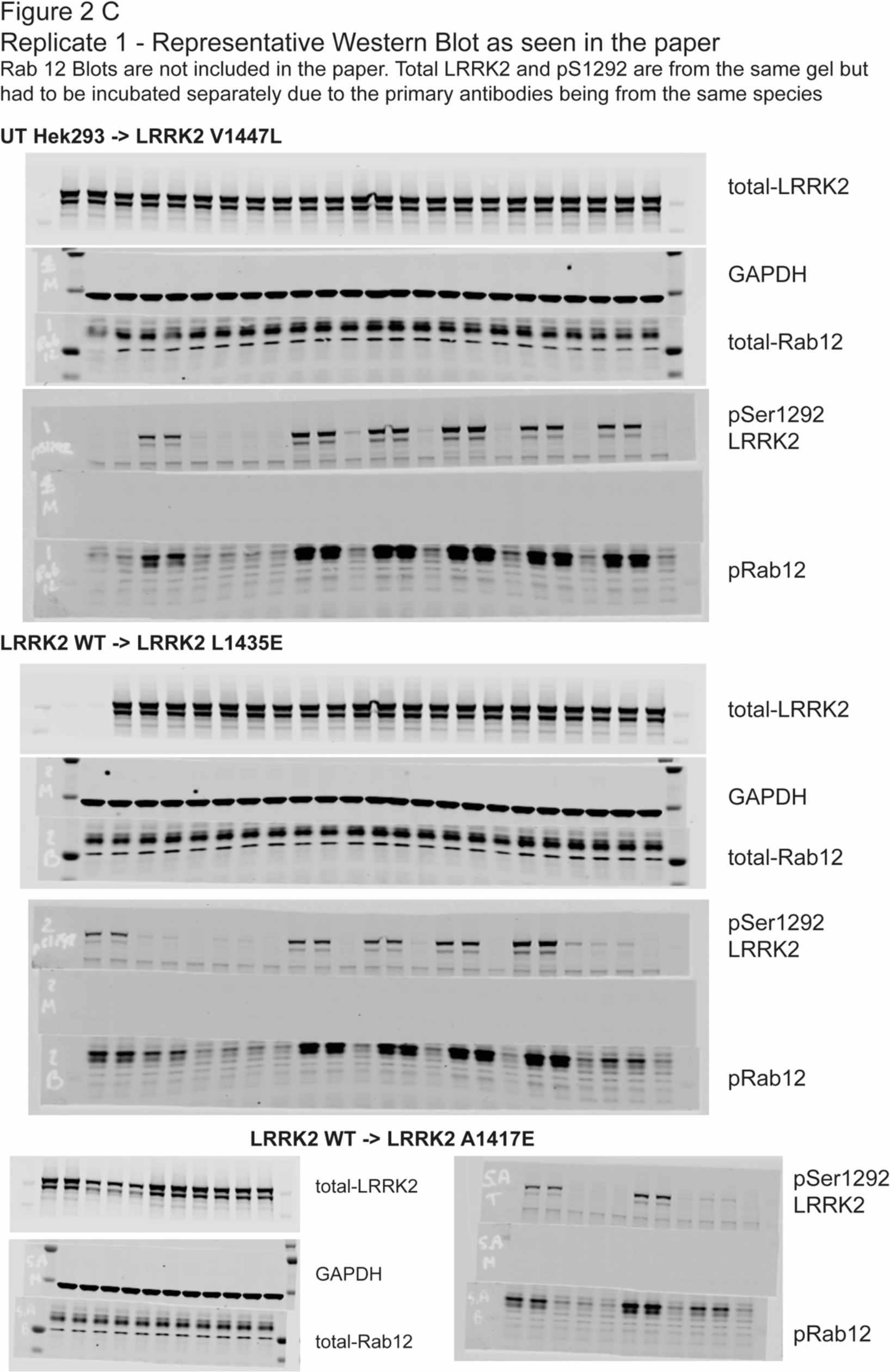
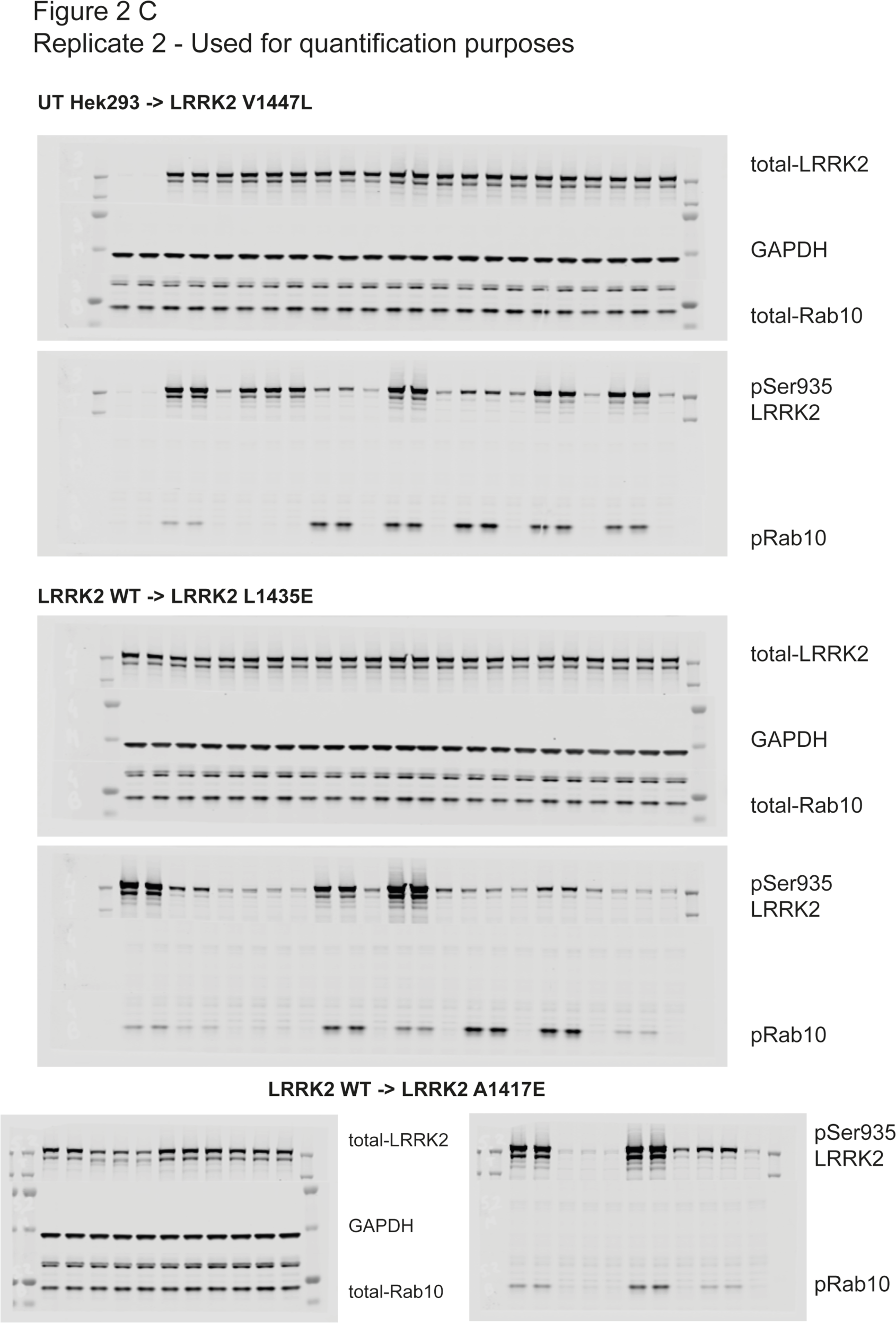
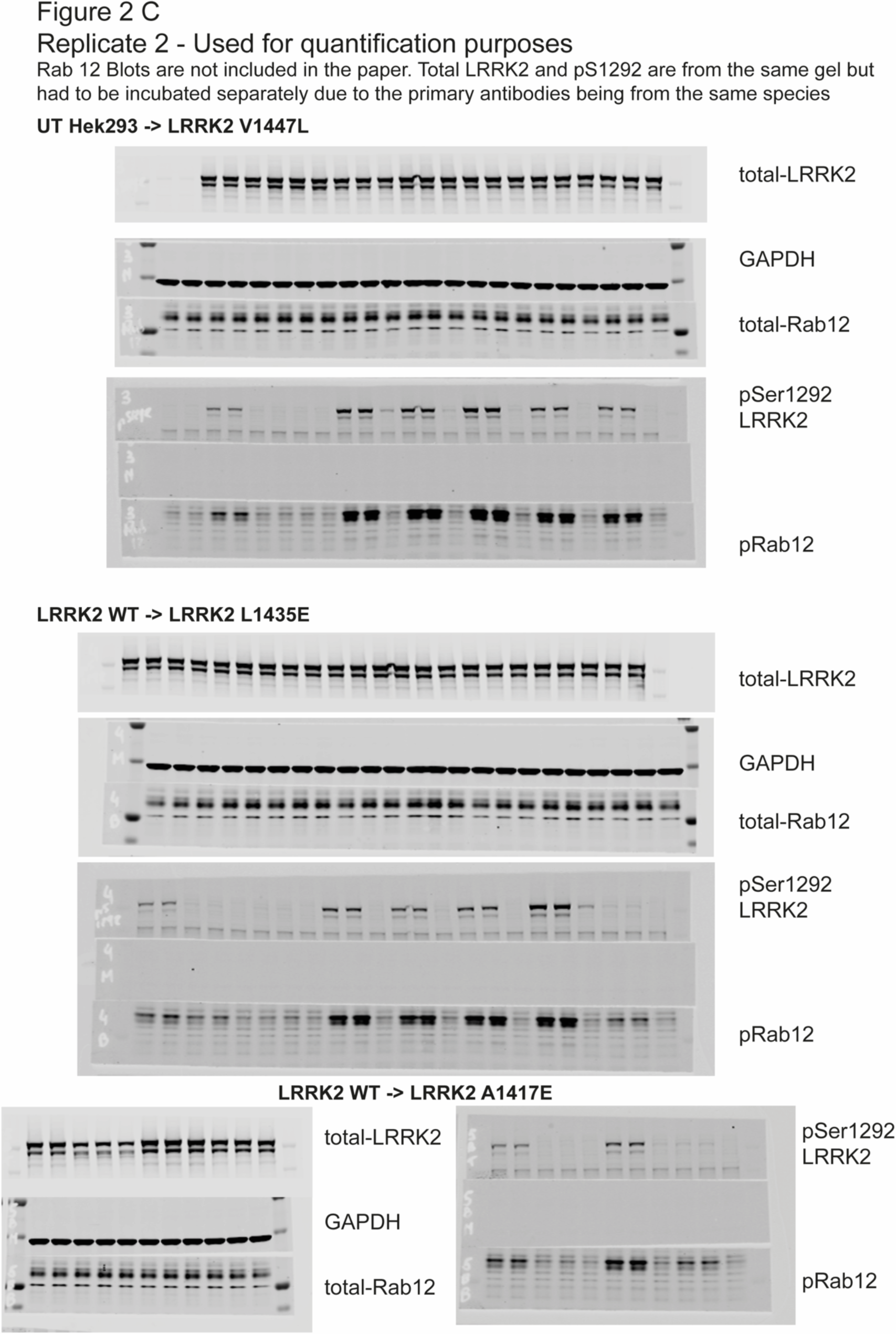
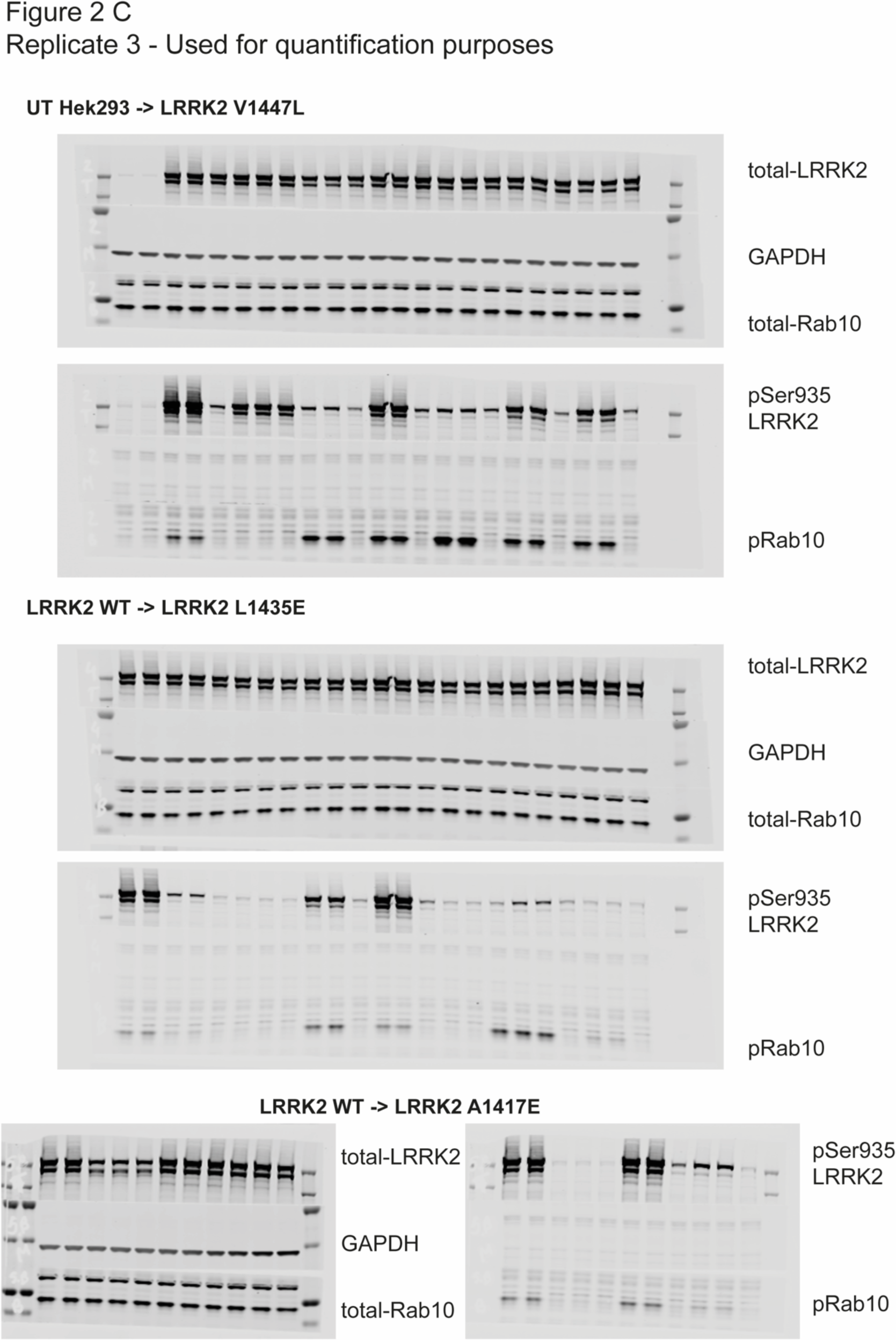
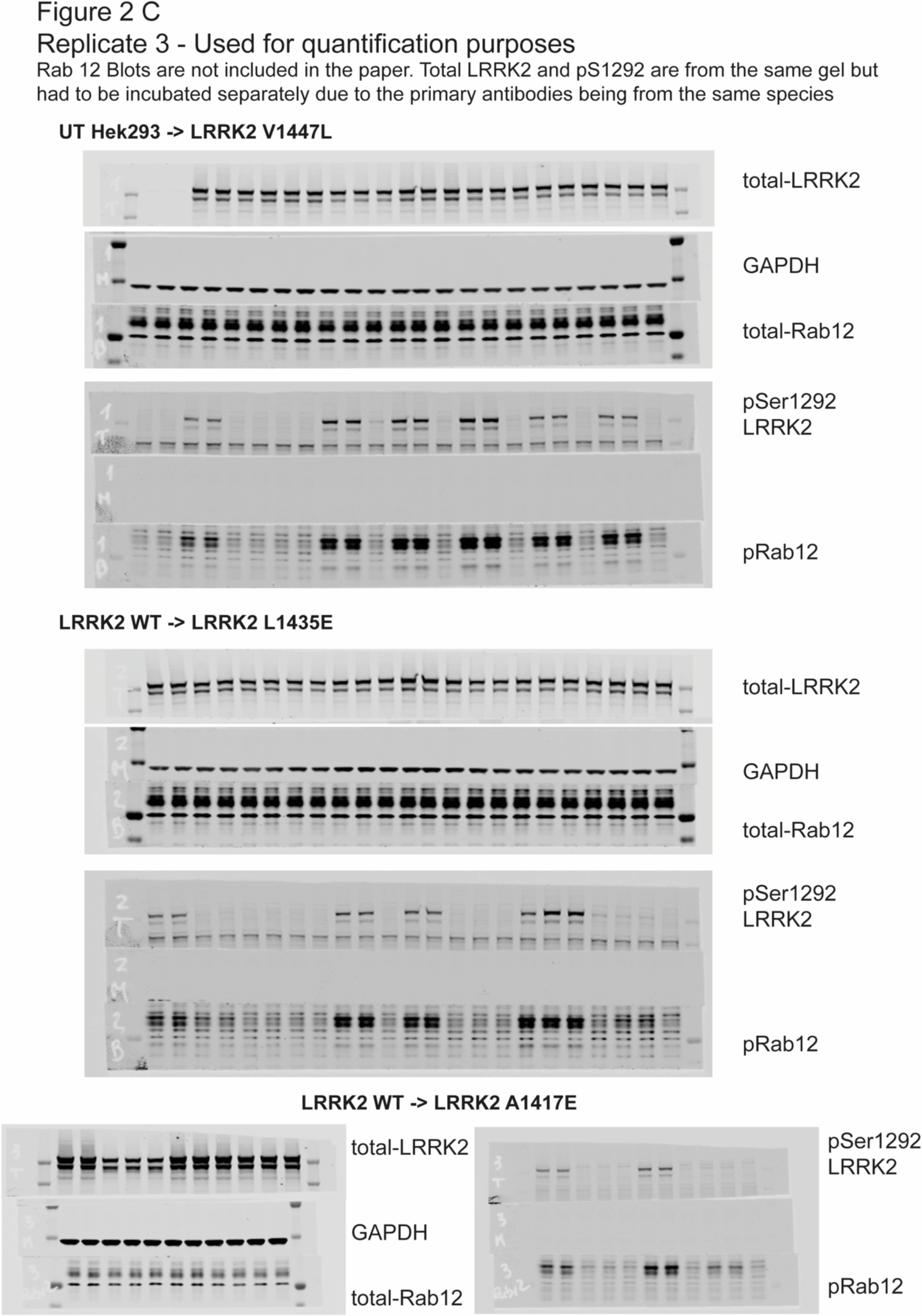

### Quantified and analysed values from immunoblotting assay as illustrated in Figure 1B

Quantification of pRab10 normalized to total Rab10 and total Rab10 normalized to GAPDH in neutrophils and monocytes from a healthy control and LRRK2 V1447L mutation carrier, treated with DMSO or MLi-2.

### Neutrophils

| pRab10/tRab10 |  |  |  | tRab10/GAPDH |  |  |
| --- | --- | --- | --- | --- | --- | --- |
|  | *DMSO* | *MLi-2* |  |  | *DMSO* | *MLi-2* |
| **Healthy control** | 1 | 0.094 |  | **Healthy control** | 1 | 0.832 |
| **LRRK2 V1447L** | 3.214 | 0.081 |  | **LRRK2 V1447L** | 1.553 | 1.711 |

### Monocytes

| pRab10/tRab10 |  |  |  | tRab10/GAPDH |  |  |
| --- | --- | --- | --- | --- | --- | --- |
|  | *DMSO* | *MLi-2* |  |  | *DMSO* | *MLi-2* |
| **Healthy control** | 1 | 0.163 |  | **Healthy control** | 1 | 0.662 |
| **LRRK2 V1447L** | 4.77 | 0.39 |  | **LRRK2 V1447L** | 0.789 | 0.39 |

### Quantified and analysed values from immunoblotting assay as illustrated in Figure 2C

**pRab10/tRab10** Quantification Across LRRK2 Mutants

Quantification of pRab10 normalized to total Rab10 in wild-type and various LRRK2 mutant under DMSO and MLi-2 treatment, with three biological replicates per condition.

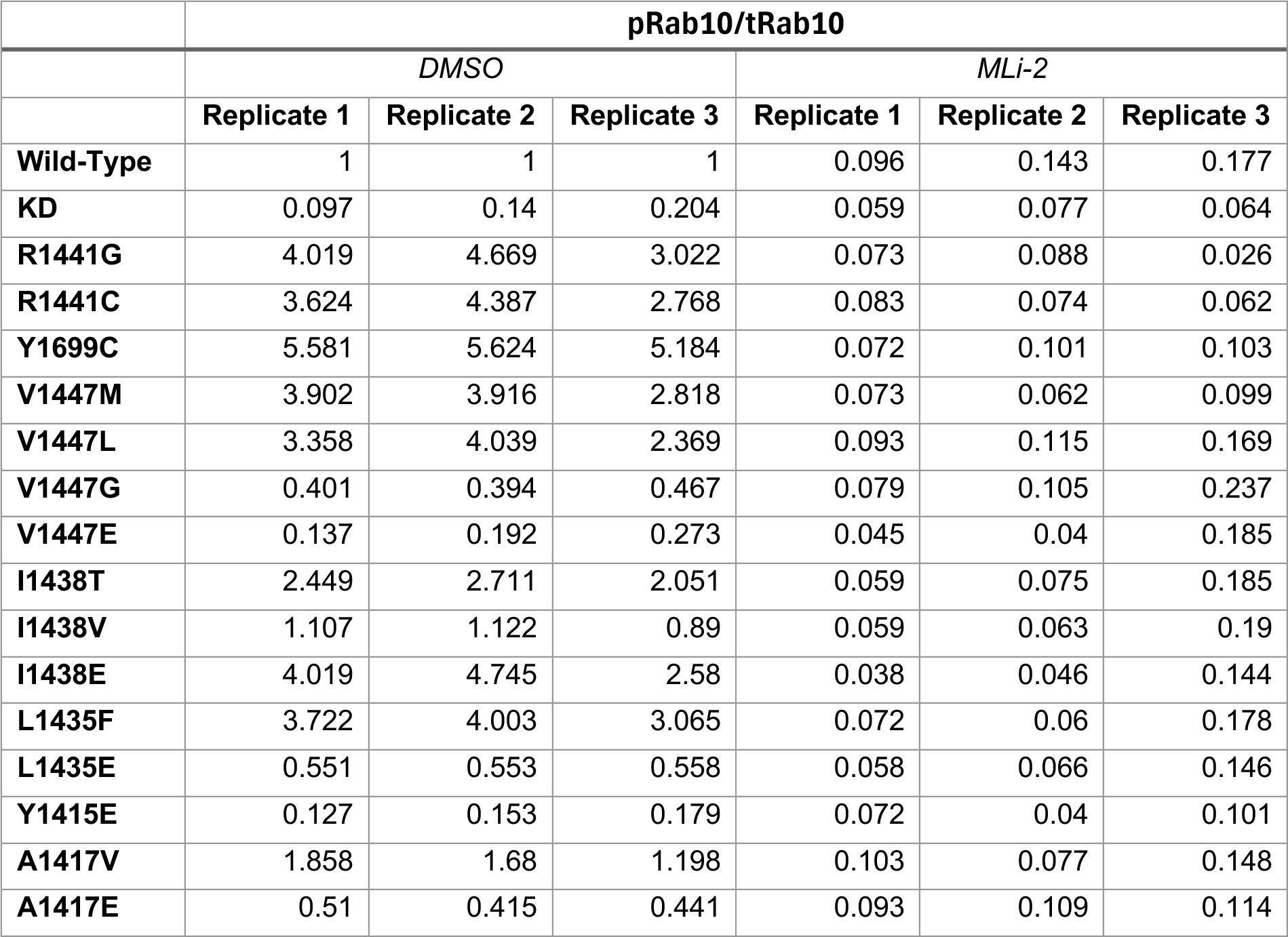

|  | **pRab10/tRab10** |  |  |  |  |  |
| --- | --- | --- | --- | --- | --- | --- |
|  | *DMSO* |  |  | *MLi-2* |  |  |
|  | **Replicate 1** | **Replicate 2** | **Replicate 3** | **Replicate 1** | **Replicate 2** | **Replicate 3** |
| **Wild-Type** | 1 | 1 | 1 | 0.096 | 0.143 | 0.177 |
| **KD** | 0.097 | 0.14 | 0.204 | 0.059 | 0.077 | 0.064 |
| **R1441G** | 4.019 | 4.669 | 3.022 | 0.073 | 0.088 | 0.026 |
| **R1441C** | 3.624 | 4.387 | 2.768 | 0.083 | 0.074 | 0.062 |
| **Y1699C** | 5.581 | 5.624 | 5.184 | 0.072 | 0.101 | 0.103 |
| **V1447M** | 3.902 | 3.916 | 2.818 | 0.073 | 0.062 | 0.099 |
| **V1447L** | 3.358 | 4.039 | 2.369 | 0.093 | 0.115 | 0.169 |
| **V1447G** | 0.401 | 0.394 | 0.467 | 0.079 | 0.105 | 0.237 |
| **V1447E** | 0.137 | 0.192 | 0.273 | 0.045 | 0.04 | 0.185 |
| **I1438T** | 2.449 | 2.711 | 2.051 | 0.059 | 0.075 | 0.185 |
| **I1438V** | 1.107 | 1.122 | 0.89 | 0.059 | 0.063 | 0.19 |
| **I1438E** | 4.019 | 4.745 | 2.58 | 0.038 | 0.046 | 0.144 |
| **L1435F** | 3.722 | 4.003 | 3.065 | 0.072 | 0.06 | 0.178 |
| **L1435E** | 0.551 | 0.553 | 0.558 | 0.058 | 0.066 | 0.146 |
| **Y1415E** | 0.127 | 0.153 | 0.179 | 0.072 | 0.04 | 0.101 |
| **A1417V** | 1.858 | 1.68 | 1.198 | 0.103 | 0.077 | 0.148 |
| **A1417E** | 0.51 | 0.415 | 0.441 | 0.093 | 0.109 | 0.114 |

**pSer1292/LRRK2** Quantification Across LRRK2 Mutants

Quantification of phosphorylated LRRK2 at Ser1292 relative to total LRRK2 in wild-type and mutant LRRK2, treated with DMSO or MLi-2, with three biological replicates per condition.

|  | **pSer1292/LRRK2** |  |  |  |  |  |
| --- | --- | --- | --- | --- | --- | --- |
|  | *DMSO* |  |  | *MLi-2* |  |  |
|  | **Replicate 1** | **Replicate 2** | **Replicate 3** | **Replicate 1** | **Replicate 2** | **Replicate 3** |
| **Wild-Type** | 1 | 1 | 1 | 0.139 | 0.161 | 0.166 |
| **KD** | 0.097 | 0.113 | 0.133 | 0.098 | 0.122 | 0.158 |
| **R1441G** | 3.046 | 3.945 | 3.116 | 0.267 | 0.527 | 0.383 |
| **R1441C** | 2.217 | 2.848 | 2.425 | 0.137 | 0.195 | 0.224 |
| **Y1699C** | 4.116 | 5.333 | 4.775 | 0.13 | 0.153 | 0.199 |
| **V1447M** | 1.743 | 1.918 | 1.432 | 0.103 | 0.07 | 0.169 |
| **V1447L** | 1.662 | 1.722 | 1.494 | 0.112 | 0.111 | 0.181 |
| **V1447G** | 0.166 | 0.213 | 0.15 | 0.061 | 0.128 | 0.099 |
| **V1447E** | 0.145 | 0.171 | 0.156 | 0.073 | 0.122 | 0.093 |
| **I1438T** | 1.494 | 1.886 | 1.322 | 0.083 | 0.122 | 0.083 |
| **I1438V** | 1.122 | 1.259 | 0.985 | 0.119 | 0.173 | 0.143 |
| **I1438E** | 1.924 | 1.926 | 1.496 | 0.067 | 0.108 | 0.148 |
| **L1435F** | 4.454 | 5.561 | 3.819 | 0.37 | 0.571 | 0.399 |
| **L1435E** | 0.241 | 0.232 | 0.286 | 0.052 | 0.101 | 0.134 |
| **Y1415E** | 0.21 | 0.23 | 0.337 | 0.164 | 0.123 | 0.22 |
| **A1417V** | 1.225 | 1.299 | 1.307 | 0.107 | 0.144 | 0.18 |
| **A1417E** | 0.239 | 0.261 | 0.248 | 0.052 | 0.098 | 0.032 |

Figure 2C – pSer935/LRRK2 Quantification Across LRRK2 Mutants Quantification of phosphorylated LRRK2 at Ser935 relative to total LRRK2 in wild-type and mutant LRRK2, treated with DMSO or MLi-2, with three biological replicates per condition.

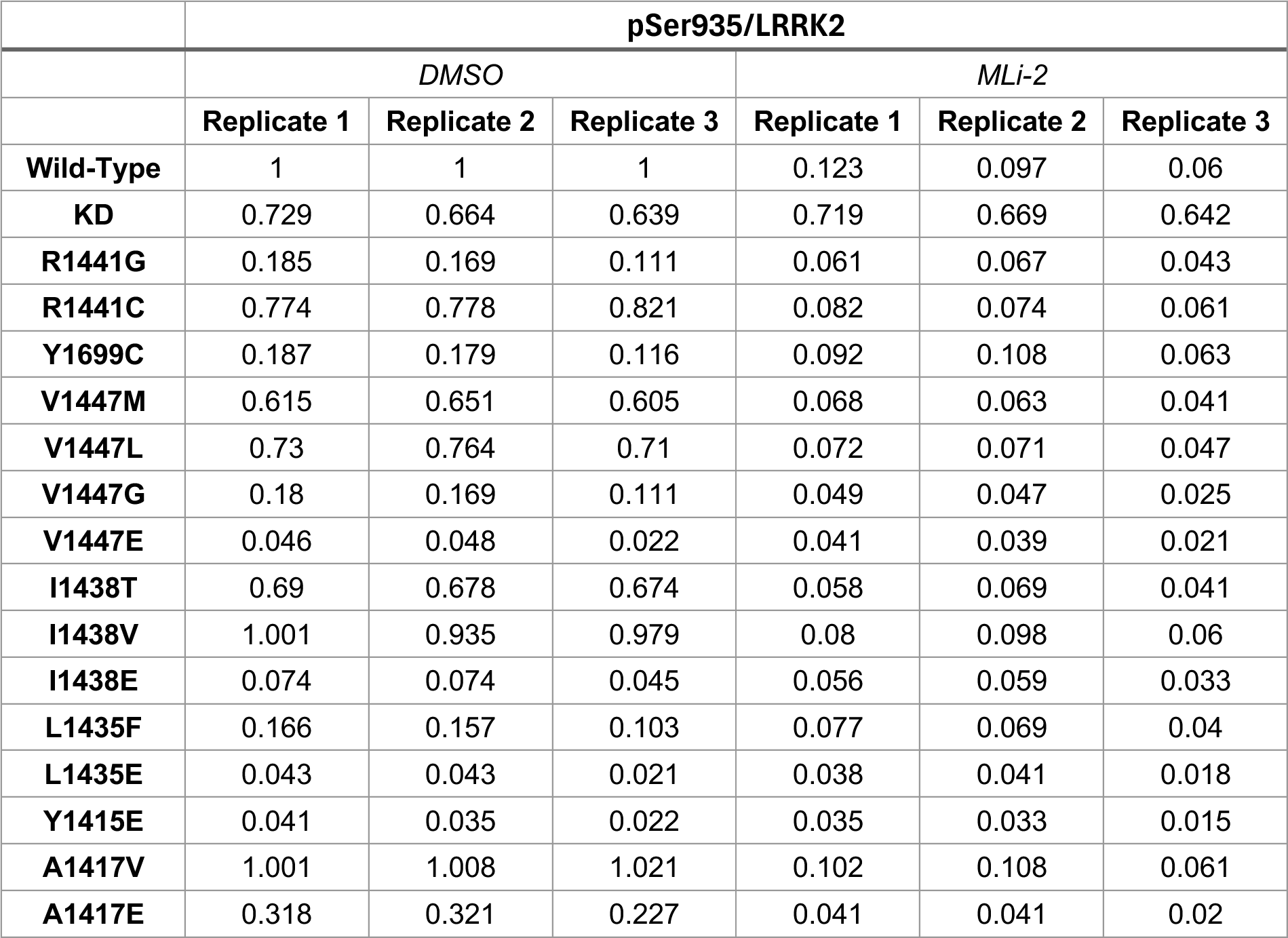

|  | **pSer935/LRRK2** |  |  |  |  |  |
| --- | --- | --- | --- | --- | --- | --- |
|  | *DMSO* |  |  | *MLi-2* |  |  |
|  | **Replicate 1** | **Replicate 2** | **Replicate 3** | **Replicate 1** | **Replicate 2** | **Replicate 3** |
| **Wild-Type** | 1 | 1 | 1 | 0.123 | 0.097 | 0.06 |
| **KD** | 0.729 | 0.664 | 0.639 | 0.719 | 0.669 | 0.642 |
| **R1441G** | 0.185 | 0.169 | 0.111 | 0.061 | 0.067 | 0.043 |
| **R1441C** | 0.774 | 0.778 | 0.821 | 0.082 | 0.074 | 0.061 |
| **Y1699C** | 0.187 | 0.179 | 0.116 | 0.092 | 0.108 | 0.063 |
| **V1447M** | 0.615 | 0.651 | 0.605 | 0.068 | 0.063 | 0.041 |
| **V1447L** | 0.73 | 0.764 | 0.71 | 0.072 | 0.071 | 0.047 |
| **V1447G** | 0.18 | 0.169 | 0.111 | 0.049 | 0.047 | 0.025 |
| **V1447E** | 0.046 | 0.048 | 0.022 | 0.041 | 0.039 | 0.021 |
| **I1438T** | 0.69 | 0.678 | 0.674 | 0.058 | 0.069 | 0.041 |
| **I1438V** | 1.001 | 0.935 | 0.979 | 0.08 | 0.098 | 0.06 |
| **I1438E** | 0.074 | 0.074 | 0.045 | 0.056 | 0.059 | 0.033 |
| **L1435F** | 0.166 | 0.157 | 0.103 | 0.077 | 0.069 | 0.04 |
| **L1435E** | 0.043 | 0.043 | 0.021 | 0.038 | 0.041 | 0.018 |
| **Y1415E** | 0.041 | 0.035 | 0.022 | 0.035 | 0.033 | 0.015 |
| **A1417V** | 1.001 | 1.008 | 1.021 | 0.102 | 0.108 | 0.061 |
| **A1417E** | 0.318 | 0.321 | 0.227 | 0.041 | 0.041 | 0.02 |

